# An Opportune and Relevant Design for Studying the Health Trajectories of Healthcare Workers

**DOI:** 10.1101/2020.06.30.20140046

**Authors:** Joseph E. Ebinger, Gregory J. Botwin, Christine M. Albert, Anders H. Berg, Aleksandra Binek, Patrick Botting, Cory Brystrom, Cindy Chavira, Jane C. Figueiredo, Soniya S. Ghandi, Caroline Lubick Goldzweig, Wohaib Hasan, Mir Henglin, Khalil Huballa, Shehnaz K. Hussain, Mohit Jain, John E. Jenrette, Sandy Joung, Elizabeth H. Kim, Angela McArdle, Shaun J. Miller, Margo Minissian, Trevor Trung Nguyen, Blandine Chazarin, Koen Raedschelders, Mohamad A. Rashid, Richard V. Riggs, Kimia Sobhani, Nancy Sun, Warren G. Tourtellotte, Dermot P. B. McGovern, Jonathan G. Braun, Jennifer E. Van Eyk, Susan Cheng, Peggy B. Miles

## Abstract

We describe the rationale, design, and implementation of baseline enrollment for a longitudinal, observational cohort study of healthcare workers. The importance of workplace as a characteristic of interest in population-based studies has become increasingly apparent over time. Most adults spend a majority of their waking hours living in their workplace environment, which serves as a source of unique as well as shared exposures, stressors, and related determinants of health. A rapidly expanding segment of the working population includes those individuals who work in the fields of healthcare. The healthcare workforce, at large, represents an increasingly diverse subset of the population with broad exposures; some exposures are specific to the workplace and others are common to persons living in the community. Importantly, healthcare workers tend to have general interest and willingness to participate in research, in addition to stable employment and easy access to on-site clinical research assessment locations. Accordingly, from a population of individuals working at a large medical delivery network, we enrolled over 6300 healthcare workers into a cohort study involving survey data collection on medical history, work environment, and family and living situation; this information was linked to participant electronic health record data and collected biospecimens.

---

The modern construct of a longitudinal observational cohort originated in the mid 20^th^ century with the founding of renowned studies such as the Framingham Heart Study(1). The design and implementation of such community-based studies have been instrumental for enabling rigorous identification of individual level characteristics associated with risk for future disease. Early and ongoing discoveries from these studies, leading to coining of the term “risk factors” (2), have had far-reaching implications for public health. The general motivation for conducting longitudinal observational studies, further exemplified in the National Health Institutes sponsored *All of Us* study (3), is centered on the premise that comprehensively evaluating exposures and outcomes can shed further light on determinants of health outcomes over the life course – for all individuals living in the community. Notwithstanding the importance of systematically observing broadly defined populations, certain subpopulations warrant special attention and, in turn, more focused investigation. Such special populations can be identified based on demographic, social, geographic, or other factors. A characteristic of particular interest – and one that identifies still understudied subgroups of individuals in the community – is workplace. In turn, an actively growing segment of our employed population with an increasingly important set of shared exposures is represented by healthcare workers. To this end, we have initiated a longitudinal cohort study that is intentionally designed to examine exposures and outcomes across a diverse population of healthcare workers living and working in Southern California.

## METHODS

### Sampling Strategy

We planned for the basis of our cohort study enrollment, along with its periodic point-of-care surveillance, to be integrated closely with the routine activities of the Employee Health Services for several reasons. First, all healthcare workers are already registered with Employee Health Services for occupational health screening, which includes a standardized assessment of medical and social history including a detailed record of vaccinations. Second, all prior and ongoing potential occupation specific exposures (e.g. airborne, bloodborne, needlestick, etc), along with the timing and nature of these exposures, are assessed and recorded by Employee Health Services. Finally, all active healthcare workers undergo annual re-assessments of vaccination status and medical history, including facilitated completion of yearly tuberculosis testing and seasonal influenza vaccination.

For our cohort study enrollment, we included all employees registered with Employee Health Services at a given healthcare facility within the network; we excluded only those employees who elected not to participate. Enrolled individuals, herein referred to as “healthcare workers” comprised the broad range of employees at our healthcare facility, including: clinical providers (physicians, advanced healthcare professionals, nurses, etc.), clinical operations and administrative staff (patient transport personnel, technicians, managerial assistants, etc.), research faculty and staff, clinical and research trainees, contractors, and hospital volunteers.

The first institution to open enrollment was the Cedars Sinai, a multi-site clinical care organization located in Los Angeles, California, that includes two major hospital centers and multiple outpatient clinics. The Cedars Sinai employee base, comprising the source population for this vanguard cohort, includes over 14,000 total employees including physicians, nursing professional, other healthcare workers who provide either direct patient care or support services, and an additional employees who are involved in primarily administrative, research, or other activities at the healthcare facility.

During the design and planning of the cohort study, the coronavirus disease 2019 (Covid-19) pandemic swept the globe (4). Given strong interest in accelerating the program in attempts to better understand Covid-19’s effect on healthcare workers and the community at large, we began study enrollment on May 11, 2020, with the support of institutional leadership and leveraging the existing Employee Health Services operational infrastructure including employee contact mechanisms and laboratory testing.

### Study enrollment and procedures

Following protocol approvals from our institutional review board, all healthcare workers employed in the medical delivery network were notified of open study enrollment via institution-wide emails, campus flyers, and announcements at regularly scheduled departmental meetings. Enrollment for each participant involved completion of an electronic informed consent form and an electronic questionnaire designed to collect data on demographic, social, and medical history characteristics (including exposures and risk factors related to Covid-19). Both the informed consent and questionnaire forms were implemented electronically via applications supported by REDCap (Research Electronic Data Capture) (5, 6). REDCap is a secure, web-based data capture tool allowing for customization of survey tools, audit and tracking of data management, automated export procedures, and data integration. Per standard clinical research procedures, all data captured by the REDCap applications are hosted on institutional servers, providing HIPPA compliant data storage and security.

Biospecimen sampling as part of the baseline protocol involved peripheral phlebotomy, conducted using standardized procedures, to enable a range of biomarker and molecular profiling assays including serology, acute phase biomarker assays, metabolomics, proteomics, and genomics. All participants were asked to provide additional opt-in consent to allow for their collected biospecimens to be included in genomic analyses. Participants were also informed regarding which results of biomarker profiling (e.g. certain serology assays performed in a CLIA certified laboratory) would be returned and which results (e.g. novel biomarker assays performed for research purposes) would not be returned. We also provided informational resources to assist with interpretation of returned results (e.g. Covid-19 serology assays) for both participants and their primary care providers.

Data management for the baseline protocol involved linking electronic questionnaire data, laboratory data, and existing electronic health record (EHR) data using the unique medical record number that was converted to a masked study identification number for statistical analyses. All data linkage processes involved initial application of data merging algorithms with output files iteratively reviewed using standard quality assurance and quality control procedures. As part of our standard data cleaning and management procedures, all identified matching issues were manually resolved by trained research staff through two-step verification of all relevant primary data sources. Data analyses for descriptive and association statistics were performed by statistically trained data analysts blinded to all participant identifiers.

All data collection, biospecimen collection, data management, and data analysis protocols and procedures were developed, reviewed, and implemented by dedicated clinical research operations, laboratory science, and data analytics teams with oversight provided by a scientific steering committee. All study protocols were approved by our institutional review boards.

## RESULTS

The initial study enrollment period occurred May 11, 2020 and included N=6318 individual employees. Demographic, social, and clinical characteristics are summarized in **Table 1**. As shown, the total study sample included 69% women, 50% non-white participants, and healthcare worker employees representing a range of age groups as well as the diversity of occupational environments relevant to the healthcare setting. Of all enrolled participants, 94.4% completed the majority (at least 90%) of all questionnaire items, 99% provided peripheral blood specimens of adequate quality for standard biomarker processing (e.g. antibody assays), and 54.8% opted in for participation in genetic analyses. As shown in **Figure 1**, the demographic characteristics of the sampled study population were similar to those of the total population of eligible study participants, without evidence of substantial under-sampling or over-sampling based on comparisons of trait frequency between the study cohort and source cohort.

**Table 1.** Sample Characteristics of the Study Cohort at Baseline.

|  | **Total** | **Intensive Care** | **Emergent or Urgent Care** | **Floor** | **Clinic** | **Office** | **Other** |
| --- | --- | --- | --- | --- | --- | --- | --- |
| n | 6318 | 1055 | 367 | 1518 | 1083 | 547 | 1748 |
| Age, mean ± SD | 41.3 (12.0) | 39.1 (10.8) | 39.0 (10.5) | 39.7 (11.2) | 42.8 (12.1) | 44.1 (13.5) | 42.9 (12.5) |
| Male, n (%) | 1987 (31) | 405 (38) | 126 (34) | 1409 (27) | 281 (26) | 177 (32) | 589 (34) |
| Hispanic ethnicity, n (%) | 1224 (19) | 136 (13) | 71 (19) | 255 (17) | 264 (24) | 135 (25) | 363 (21) |
| Race (%) |  |  |  |  |  |  |  |
| African American/Black | 391 (6) | 67 (6) | 24 (7) | 107 (7) | 60 (6) | 33 (6) | 100 (6) |
| American Indian/Alaska Native | 18 (0) | 3 (0) | 4 (1) | 5 (0) | 1 (0) | 1 (0) | 4 (0) |
| Asian | 1953 (31) | 369 (35) | 82 (22) | 569 (38) | 234 (22) | 131 (24) | 568 (33) |
| Pacific Islander | 59 (1) | 8 (1) | 5 (1) | 19 (1) | 9 (1) | 4 (1) | 14 (1) |
| White | 3138 (50) | 485 (46) | 184 (50) | 631 (42) | 642 (59) | 311 (57) | 885 (51) |
| Other | 759 (12) | 123 (12) | 68 (19) | 187 (12) | 137 (13) | 67 (12) | 177 (10) |
| Medical Conditions, n (%) |  |  |  |  |  |  |  |
| Asthma | 747 (12) | 128 (12) | 48 (13) | 175 (12) | 152 (14) | 78 (14) | 166 (10) |
| Autoimmune disease | 227 (4) | 43 (4) | 13 (4) | 42 (3) | 44(4) | 26 (5) | 64 (4) |
| Cancer | 190 (3) | 31 (3) | 9 (3) | 31 (2) | 35 (3) | 19 (3) | 65 (4) |
| Chronic obstructive pulmonary disease | 83 (1) | 14 (1) | 4 (1) | 14 (1) | 17 (2) | 6 (1) | 28 (2) |
| Coronary disease or heart failure | 124 (2) | 17 (2) | 5 (1) | 24 (2) | 26 (2) | 14 (2.6) | 38 (2) |
| Diabetes | 377 (6) | 57(5) | 18 (5) | 89 (6) | 67(6) | 30 (5.5) | 116 (7) |
| Hypertension | 987 (16) | 152 (14) | 51 (14) | 216 (14) | 192 (18) | 104 (19) | 272 (16) |
| Marital Status, n (%) |  |  |  |  |  |  |  |
| Married | 2939 (47) | 460 (44) | 166 (45) | 706 (47) | 579 (54) | 277 (51) | 751 (43) |
| Single | 2075 (33) | 429 (41) | 133 (36) | 557 (37) | 315 (29) | 173 (32) | 468 (27) |
| Other | 1304 (21) | 166 (16) | 68 (19) | 255 (17) | 189 (17) | 97 (18) | 529 (30) |
| Current Smoking, n (%) | 105 (2) | 18 (2) | 4 (1) | 32 (2) | 12 (1) | 7 (1) | 32 (2) |
| Night Shift, n (%) | 1151 (18) | 382 (36) | 121 (33) | 454 (30) | 14 (1) | 11 (2) | 169 (10) |
| Rotating Shifts, n (%) | 729 (12) | 253 (24) | 79 (22) | 212 (14) | 31 (3) | 8 (2) | 146 (8) |
| Work Hours/Week | 40.8 (11.1) | 42.9 (13.5) | 37.6 (10.1) | 40.5 (12.3) | 41.2 (9.2) | 40.9 (8.5) | 40.1 (9.8) |

| Dwelling Type, n (%) |  |  |  |  |  |  |  |
| --- | --- | --- | --- | --- | --- | --- | --- |
| Apartment | 2763 (44) | 528 (50) | 193 (53) | 721 (48) | 425 (39) | 242 (44) | 654 (37) |
| House | 3053 (48) | 484 (46) | 157 (43) | 732 (48) | 623 (58) | 287 (53) | 770 (44) |
| Other | 502 (8) | 43 (4) | 17 (5) | 65 (4) | 35 (3) | 18 (3) | 324 (19) |
| No. People in the Home | 2.3 (1.7) | 2.0 (1.7) | 2.2 (1.7) | 2.4 (1.7) | 2.4 (1.7) | 2.3 (1.7) | 2.3 (1.7) |

Values are shown as or mean±SD for continuous variables or number (percent) frequency for categorical variables.

**Figure 1.**
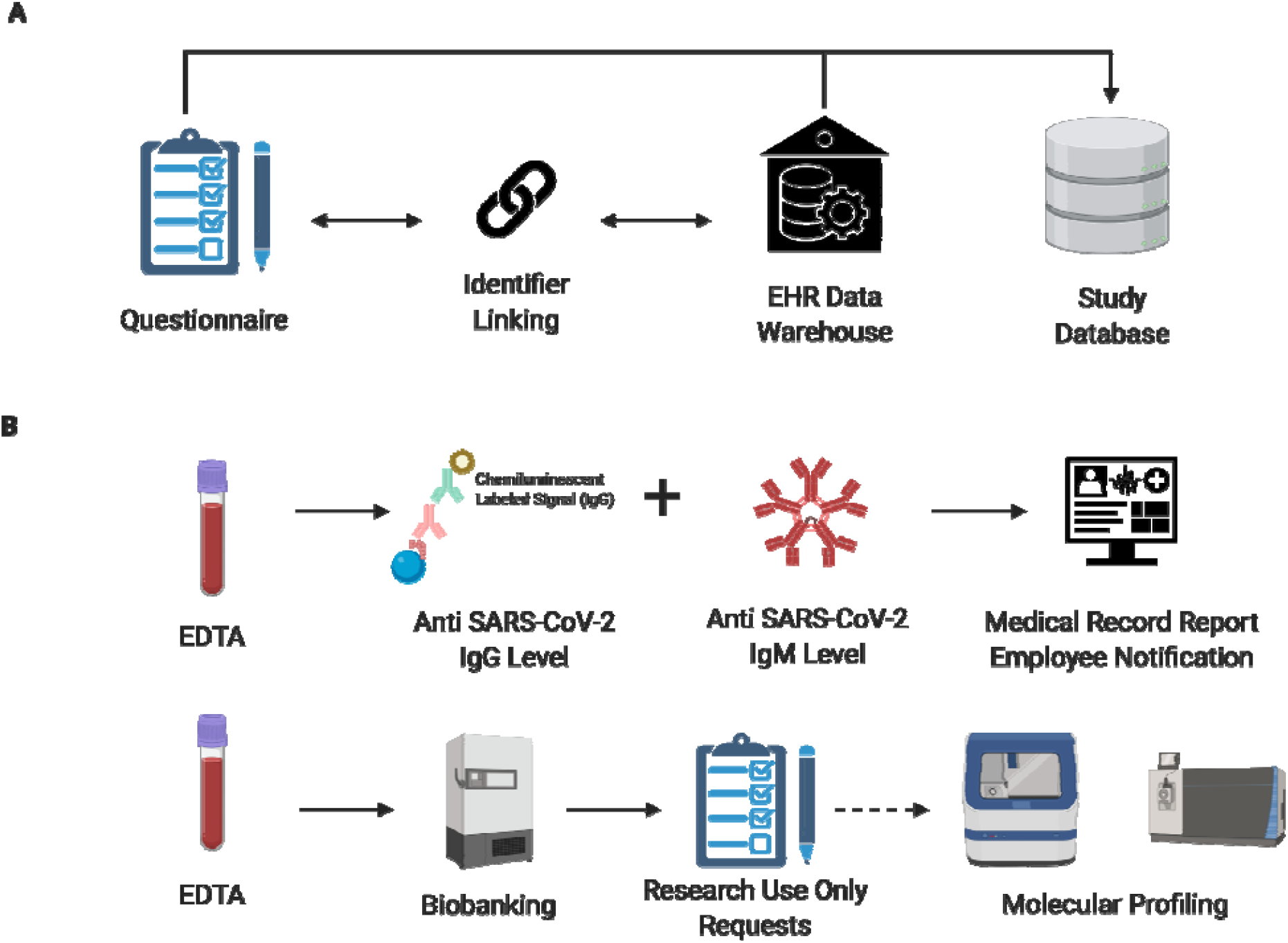
Baseline Enrollment Process. The general data collection workflow is shown **Panel A**. The general biospecimen collection workflow is shown in **Panel B**.

**Figure 2.**
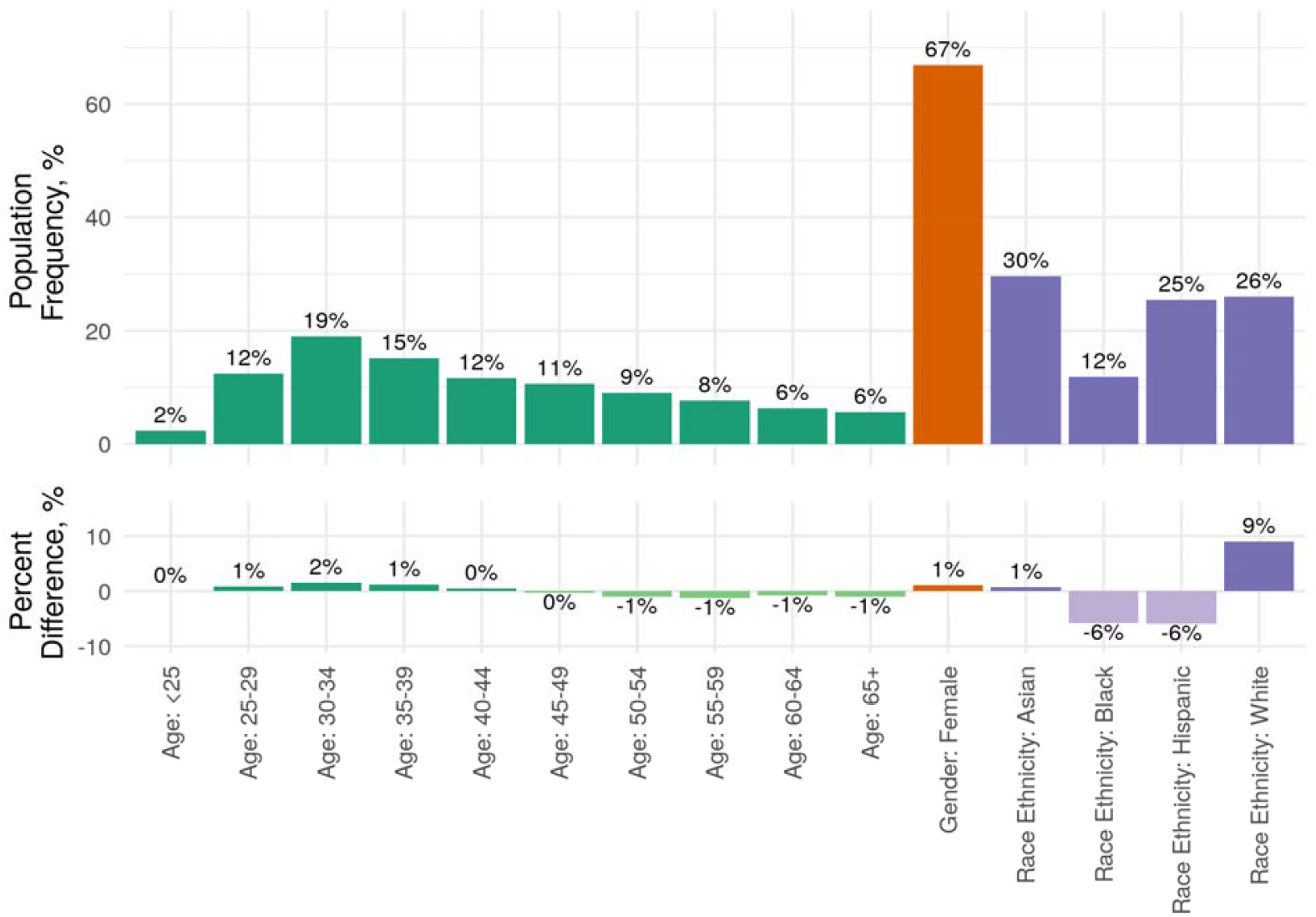
Overview of Sampling Selection. Source population level demographics are shown in the upper panel, and the difference in proportion of sampled participants compared to the source population is shown in the lower panel for each demographic trait.

## DISCUSSION

We have designed and implemented a healthcare worker cohort study that has successfully enrolled over 6300 participants including women and men across a diversity of age groups, racial and ethnic backgrounds, and occupational environments. In addition to diverse enrollment, representing a large proportion of the source population, we found that participants were engaged in active recruitment over a relatively short period of time, with a high rate of questionnaire completion and a majority of participants opting in for genetic testing. We recognize that efficient enrollment and effective engagement could be achieved with focused recruitment communications, making certain clinically relevant information returnable to participants (e.g. antibody test results), and leveraging an existing infrastructure to accommodate a high volume of participant interest.

Our healthcare worker cohort study expands from the design of prior studies by facilitating engagement in-person through an Employee Health Services infrastructure as well as remotely via electronic and mail communications (7-14). Similar to prior cohort studies of healthcare workers, the current study is designed to support investigations that are both specific to exposures enriched among persons working in healthcare settings and also generalizable to exposures shared across communities at large. Similar to other cohorts, the size of the current study is intentionally large so that future ancillary studies may include embedded clinical trials of interventions with adequate statistical power. As an extension from prior work, the current cohort was recruited from a medical delivery network based in the populous metropolitan region of Los Angeles County in Southern California; this regional base has allowed sampling of participants representing a spectrum of racial, ethnic, socioeconomic, and educational backgrounds that can be considered diverse even among healthcare workers.

A cohort of healthcare workers, such as this one, offers several operational advantages when considering future investigations. As demonstrated in prior studies with high rates of longitudinal follow up,(7, 8) healthcare workers are frequently well-versed in the importance of participating in observational studies with the potential to generate discoveries that can inform clinical practice. For the current study, with enrollment and engagement activities occurring at the place of employment, future additional in-person assessments including biospecimen collections are likely to have high adherence rates given the convenience of location. Both the workplace location and the infrastructure provided by Employee Health Services also allow for efficient methods for re-contacting participants and tracking outcomes, particularly given the integration of employee health records within a comprehensive medical records system that permits real-time verification of self-reported clinical data for those participants who also receive personal medical care within the same healthcare system.

Several limitations of the study merit consideration. Although healthcare workers tend to represent a relatively stable workforce based population, some enrollees will transfer in or out of the system over time; as has been done for similar cohort studies, we plan to retain follow-up for as many base enrollees as possible while also allowing for new participants to be enrolled in pre-specified contexts (e.g. as part of ancillary studies, with appropriate scientific justification and ability to statistically accommodate for differences in timing of measurable exposures and outcomes). While relatively diverse in composition, we recognize that our base cohort does not completely represent all members of the community at large. For example, healthcare workers tend to be physically able-bodied in order to carry out job related tasks, they are able to speak English as a primary language in the workplace, and they are required to have documented residency status. Further, given that our healthcare workers are largely employed and not retired, they tend to younger than the overall population, particularly when compared to the population of individuals with multiple chronic diseases. Healthcare workers, compared to a more general sample from the population, may also tend to have higher levels of health literacy and potentially a greater willingness to respond to invitations to participate in research and disease prevention activities.

The healthcare industry continues to grow in size and diversity, employing a rapidly expanding segment of the total working population (15). Importantly, most adults spend a majority of their waking hours within their workplace environment. Thus, investigating occupation specific as well as more general exposures among our healthcare workers promises to shed light on disease determinants that will likely be of increasing relevance to the health of the public at large. Notwithstanding the benefits of published research in this field to public health, we expect that healthcare organizations will remain actively interested in studying the health of its workers for reasons related to institutional well-being. Accordingly, given the importance of workforce health to institutions in general, we anticipate that organizations in other industries will also continue to share in this interest.

## Data Availability

The data garnered as a result of this study are available from Cedars-Sinai Medical Center, upon reasonable request. The data are not publicly available due to the contents including information that could compromise research participant privacy/consent.

## ACKNOWLEDGEMENTS

Author affiliations: Department of Cardiology, Smidt Heart Institute, Cedars Sinai Medical Center, Los Angeles, California (Joseph E. Ebinger; Christine M. Albert; Patrick Botting; Cory Bystorm; Mir Henglin; Sandy Joung; Elizabeth H. Kim; Margo Minissian; Trevor Trung Nguyen; Blandine Chazarin; Koen Raedschelders; Mohamad A. Rashid; Nancy Sun; Jennifer E. Van Eyk; Susan Cheng). F. Widjaja Foundation Inflammatory Bowel and Immunobiology Research Institute, Cedars Sinai Medical Center, Los Angeles, California (Gregory J. Botwin; Dermot P.B. McGovern; Jonathan G. Braun). Pathology and Laboratory Medicine, Cedars Sinai Medical Center, Los Angeles, California (Anders H. Berg; Wohaib Hasan; Khalil Huballa; Kimia Sobhani; Warren G. Tourtellotte). Advanced Clinical Biosystems Institute, Department of Biomedical Sciences, Cedars Sinai Medical Center, Los Angeles, California (Aleksandra Binek; Angela McArdle; Blandine Chazarin; Koen Raedschelders; Jennifer E. Van Eyk). Precision Biomarker Laboratories, Cedars Sinai Medical Center, Los Angeles, California (Cory Bystorm). Department of Medicine, Cedars Sinai Medical Center, Los Angeles, California (Cindy Chavira). Cedars-Sinai Cancer and Department of Medicine, Cedars-Sinai Medical Center, Los Angeles California (Jane C. Figueiredo; Shehnaz K. Hussain). Department of Medicine School of Medicine, University of California, San Diego, San Diego, CA (Mohit Jane). Office of Infection Control, Marina Del Rey Hospital, Marina Del Rey, California (Soniya S. Gandi). Cedars Sinai Medical Network, Cedars Sinai Medical Center, Los Angeles, California (Caroline Lubick Goldzweig; John E. Jenrette). Biobank & Translational Research Core Laboratory, Samuel Oschin Comprehensive Cancer Institute, Cedars Sinai Medical Center, Los Angeles, California (Wohaib Hasan; Warren G. Tourtellotte). Enterprise Information Services, Cedars Sinai Medical Center, Los Angeles, California (Shaun J. Miller). Brawerman Nursing Institute, Cedars Sinai Medical Center, Los Angeles, California (Margo Minissian). Chief Medical Officer, Cedars-Sinai, Los Angeles, California (Richard V. Riggs).

Joseph E. Ebinger, Gregory J. Botwin, Susan Cheng, and Peggy B. Miles all contributed equally to the work.

We thank Tessa Calahan, MBA, RD, Denica Moodley, MPH, and Christopher Simon, MBA, MSHS for all their invaluable assistance with this project. We are grateful to all the healthcare workers in our healthcare system who continue to be dedicated to delivering the highest quality care for all patients.

## Conflict of interest

none declated.

## Notes

### Competing Interest Statement

The authors have declared no competing interest.

### Funding Statement

TBD

### Author Declarations

All study protocols were approved by the Cedars-Sinai Institutional Review Board. Informed consent was obtained when required for participation.

## REFERENCES

1. Mahmood SS, Levy D, Vasan RS, et al. The Framingham Heart Study and the epidemiology of cardiovascular disease: a historical perspective. Lancet 2014;383(9921):999–1008.

2. Kannel WB, Dawber TR, Kagan A, et al. Factors of risk in the development of coronary heart disease--six year follow-up experience. The Framingham Study. Ann Intern Med 1961;55:33–50.

3. The “All of Us” Research Program. New England Journal of Medicine 2019;381(7):668–76.

4. Omer SB, Malani P, Del Rio C. The COVID-19 Pandemic in the US: A Clinical Update. JAMA 2020.

5. Harris PA, Taylor R, Thielke R, et al. Research electronic data capture (REDCap)--a metadata-driven methodology and workflow process for providing translational research informatics support. J Biomed Inform 2009;42(2):377–81.

6. Harris PA, Taylor R, Minor BL, et al. The REDCap consortium: Building an international community of software platform partners. J Biomed Inform 2019;95:103208.

7. Buring JE, Hennekens CH. Cost and efficiency in clinical trials: the U.S. Physicians’ Health Study. Stat Med 1990;9(1-2):29–33.

8. Colditz GA, Manson JE, Hankinson SE. The Nurses’ Health Study: 20-year contribution to the understanding of health among women. J Womens Health 1997;6(1):49–62.

9. Christen WG, Gaziano JM, Hennekens CH. Design of Physicians’ Health Study II--a randomized trial of beta-carotene, vitamins E and C, and multivitamins, in prevention of cancer, cardiovascular disease, and eye disease, and review of results of completed trials. Ann Epidemiol 2000;10(2):125–34.

10. Draper H, Wilson S, Ives J, et al. Healthcare workers’ attitudes towards working during pandemic influenza: a multi method study. BMC Public Health 2008;8:192.

11. Derycke H, Clays E, Vlerick P, et al. Perceived work ability and turnover intentions: a prospective study among Belgian healthcare workers. J Adv Nurs 2012;68(7):1556–66.

12. Gorar ZA, Butt ZA, Aziz I. Risk factors for bloodborne viral hepatitis in healthcare workers of Pakistan: a population based case-control study. BMJ Open 2014;4(7):e004767.

13. Post JM, Ali SS, Roberson LL, et al. Rationale and design of the Baptist Employee Healthy Heart Study: a randomized trial assessing the efficacy of the addition of an interactive, personalized, web-based, lifestyle intervention tool to an existing health information web platform in a high-risk employee population. Trials 2016;17(1):308.

14. Stampfer M. New insights from the British doctors study. BMJ 2004;328(7455):1507.

15. Moore KD, Eyestone KM, Coddington DC. Viewing U.S. health care as a growth industry. Healthc Financ Manage 2014;68(4):102, 4.

